# Systematic review and meta-analysis of randomized trials of hydroxychloroquine for the prevention of COVID-19

**DOI:** 10.1101/2020.09.29.20203869

**Authors:** Xabier García-Albéniz, Julia del Amo, Rosa Polo, José Miguel Morales-Asencio, Miguel A Hernán

## Abstract

**Background:** Recruitment into randomized trials of hydroxychloroquine (HCQ) for prevention of COVID-19 has been adversely affected by a widespread conviction that HCQ is not effective for prevention. In the absence of an updated systematic review, we conducted a meta-analysis of randomized trials that study the effectiveness of HCQ to prevent COVID-19.

**Methods:** A search of PubMed and medRxiv with expert consultation found ten completed randomized trials: seven pre-exposure prophylaxis trials and three post-exposure prophylaxis trials. We obtained or calculated the risk ratio of COVID-19 diagnosis for assignment to HCQ versus no HCQ (either placebo or usual care) for each trial, and then pooled the risk ratio estimates.

**Results:** The pooled risk ratio estimate of the pre-exposure prophylaxis trials was 0.72 (95% CI: 0.58-0.91) when using either a fixed effect or a standard random effects approach, and 0.72 (95% CI: 0.52-1.00) when using a conservative modification of the Hartung-Knapp random effects approach. The corresponding estimates for the post-exposure prophylaxis trials were 0.91 (95% CI: 0.71-1.16) and 0.91 (95% CI: 0.54-1.55). All trials found a similar rate of serious adverse effects in the HCQ and no HCQ groups.

**Discussion:** A benefit of HCQ as prophylaxis for COVID-19 cannot be ruled out based on the available evidence from randomized trials. However, the “not statistically significant” findings from early prophylaxis trials were widely interpreted as definite evidence of lack of effectiveness of HCQ. This interpretation disrupted the timely completion of the remaining trials and thus the generation of precise estimates for pandemic management before the development of vaccines.

## Background

Hydroxychloroquine (HCQ) is not an effective treatment for established coronavirus disease 2019 (COVID-19) (1, 2), but it is unclear whether HCQ can prevent symptomatic COVID-19. Early in the SARS-CoV-2 pandemic, about 30 randomized trials were designed to study HCQ as prophylaxis for COVID-19 (3). After the findings from two of these trials were reported in the Summer of 2020 (4, 5), HCQ was generally viewed by the medical community as ineffective for COVID-19 prophylaxis. The emergence of that consensus was surprising because both trials found a lower risk of COVID-19 in the HCQ group, though they were too small to rule out either benefit or harm of HCQ.

A timely completion of the remaining trials would have generated precise estimates of the potential effectiveness of HCQ to prevent COVID-19 among those at high risk of infection or complications. However, the widespread conviction about HCQ’s lack of effectiveness dramatically slowed down the recruitment into ongoing trials of HCQ prophylaxis (one of them carried out by the authors of this report) (6). As a result, key decisions were made based on insufficient evidence during the pre-vaccine period of the pandemic.

We conducted a systematic review and meta-analysis of randomized trials that study the effectiveness of HCQ to prevent COVID-19 either before known exposure to an infected individual (pre-exposure prophylaxis) or after known exposure to an infected individual (post-exposure prophylaxis).

## Methods

Studies were eligible for inclusion if they were randomized clinical trials comparing hydroxychloroquine as prophylaxis (pre-exposure or post-exposure) for COVID-19 with a non-active control, included individuals who had a polymerase chain reaction (PCR) negative test for SARS-CoV-2 at the time of treatment assignment, and had the full text published in a peer-reviewed journal or as a pre-print. Given the relatively small number of trials, we identified candidate studies through expert consultation and confirmed the findings with a search of PubMed and medRxiv as of December 8, 2021, using the search strategy described in the Appendix (first reported as part of a preprint on September 29, 2020). Two authors (XGA and MAH) independently reviewed the full text of the identified studies and extracted the data. Disagreements were resolved by consulting other co-authors (JdA, RP). Incomplete or unpublished data were requested from the investigators of each trial. The risk of bias of the included studies was assessed independently by 2 authors (XGA, MAH) using the “Rob 2” tool by the Cochrane Bias Methods Group (7).

For each of the identified trials, we obtained or calculated the risk ratio of COVID-19 for assignment to HCQ versus no HCQ (either placebo or usual care) among PCR-negative individuals at baseline. Our primary analysis is based on the definition of COVID-19 reported in the primary analysis of each study. In addition, we tried to harmonize the definition of COVID-19 across studies by conducting a separate meta-analysis for laboratory-confirmed symptomatic COVID-19.

We calculated the pooled risk ratio estimate and its 95% confidence or compatibility interval (CI) using a fixed (or common) effect approach and two types of random effects approaches (8, 9). Because the standard random effects method may yield an anticonservative (that is, too narrow) 95% CI (8), we also used the Hartung-Knapp random effects method (9), which has been shown to generally outperform the standard random effects method (10). However, this latter method results in an even more anticonservative 95% CI (narrower than the 95% CI from the standard method) when, as in our meta-analysis, the estimated between-study heterogeneity is small (tau-squared near zero) (11, 12). We therefore used the Hartung-Knapp method with an ad hoc modification (13) designed to ensure that its 95% CI remains wider than that of the standard method, even though the resulting 95% CI is expected to be conservative (too wide) when, as in our meta-analysis, the number of studies is small (14). All analyses were conducted in Stata (version 16.1; Stata Corporation, College Station, Texas, USA).

## Results

Ten completed randomized trials were identified and included in the meta-analysis: seven trials studied hydroxychloroquine as pre-exposure prophylaxis (15-21) and three as post-exposure prophylaxis (4, 5, 22).

The Table summarizes the key design characteristics and effect estimates reported by each study. The primary outcome definition varied across trials, with some trials using the presence of symptoms with or without laboratory confirmation (4, 15, 17), others using laboratory-confirmed symptomatic COVID-19 (5, 19) and others laboratory-confirmed SARS-CoV-2 infection with or without symptoms (16, 18, 22). Our risk of bias assessment identified the handling of incomplete ascertainment of the outcome and the exclusion of patients after randomization as possible sources of moderate bias (see Supplementary Material). One study (20) with moderate risk of bias was not included in the meta-analysis because the effect estimate was reported only as a p-value. Another study (21) was not included because there were zero events in the HCQ arm.

The five pre-exposure prophylaxis trials were double-blinded, placebo-controlled trials that included healthcare workers with ongoing exposure to patients with COVID-19 (15-19). The occurrence of COVID-19 was ascertained during a period between 4 and 12 weeks. Figure 1 shows the risk ratio estimates from the five pre-exposure prophylaxis trials. The pooled risk ratio of COVID-19 for hydroxychloroquine vs. no hydroxychloroquine was 0.72 (95% CI: 0.58, 0.91) when using either a fixed effect or a standard random effects approach, and 0.72 (95% CI: 0.58, 1.00) when using the ad hoc modification of the Hartung-Knapp approach. The corresponding pooled risk difference was -2.4 cases per 100 individuals (95% CI -4.3, - 0.6) when using a fixed effect, -2.7 cases per 100 individuals (95% CI -5.3, -0.2) when using a standard random effect approach and -2.7 cases per 100 individuals (95% CI -6.8, 1.34) when using the ad hoc modification of the Hartung-Knapp approach (Supplementary Figure 1). The pooled risk ratio estimate ranged from 0.71 to 0.73 under the alternative outcome definition and in a sensitivity analysis that used only studies published in peer-reviewed journals (15, 16, 18), but the 95% CIs were wider (Supplementary Figures 2-3).

**Figure 1.**
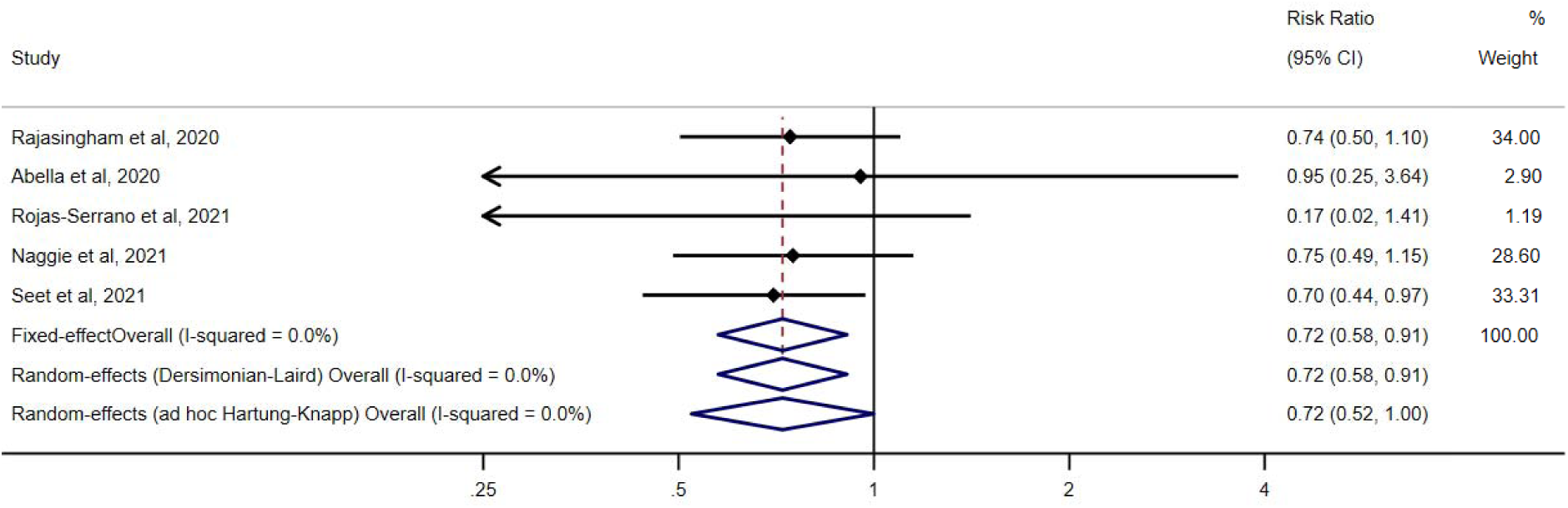
Risk ratio estimates of COVID-19 for hydroxychloroquine vs. no hydroxychloroquine in randomized trials of pre-exposure prophylaxis, pooled and by study. These estimates are based on the definition of COVID-19 reported in the primary analysis of each study.

The three post-exposure prophylaxis trials included asymptomatic contacts of confirmed COVID-19 cases. The time from exposure to initiation of prophylaxis was relatively long: in one trial, about a third of participants were enrolled 4 days after exposure (none was enrolled later) (4); in the other, about 10% of participants were enrolled 7 or more days after exposure (5); in the third study, 8% of participants received a first dose 5 days or later after exposure (22). For comparison, post-exposure prophylaxis for HIV is recommended in the first 6-72 hours after the exposure (23, 24). The occurrence of COVID-19 was ascertained during a period of 2 weeks. The pooled risk ratio of COVID-19 for assignment to hydroxychloroquine vs. no hydroxychloroquine was 0.91 (95% CI: 0.71, 1.16) when using either a fixed effect or a standard random effects approach, and 0.91 (95% CI: 0.54, 1.55) when using the ad hoc modification of the Hartung-Knapp approach (Figure 2).

**Figure 2.**
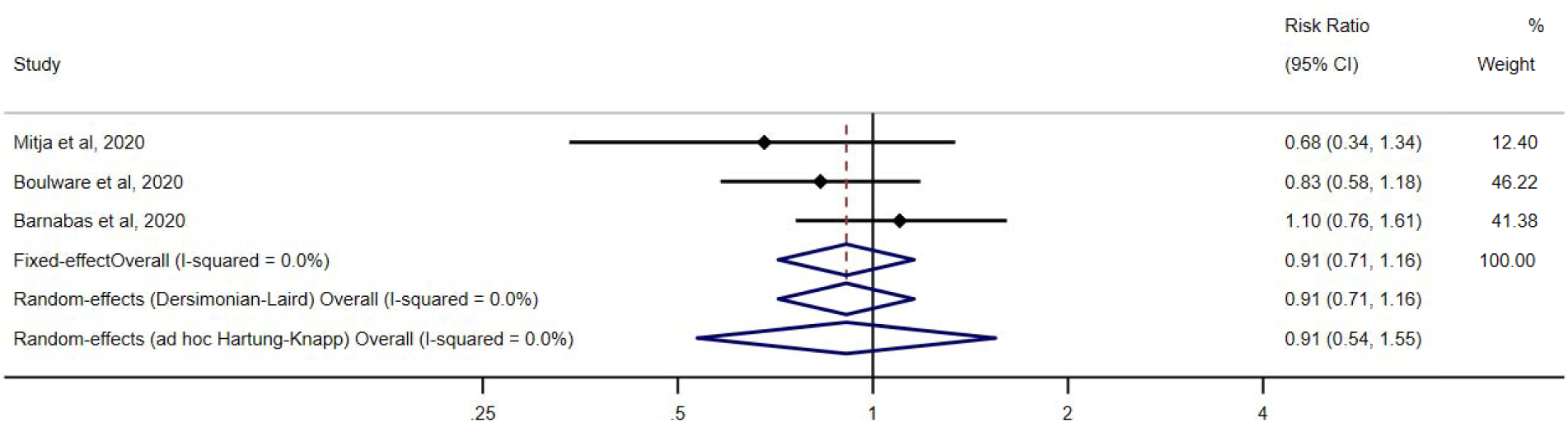
Risk ratio estimates of COVID-19 for hydroxychloroquine vs. no hydroxychloroquine in randomized trials of post-exposure prophylaxis, pooled and by study. These estimates are based on the definition of COVID-19 reported in the primary analysis of each study.

All trials found a similarly low rate of serious adverse effects in the HCQ and no HCQ groups. As expected, the proportion of mild gastrointestinal side effects was greater in the HCQ group. Adherence to treatment was heterogeneous across trials (Table)

**Table.**
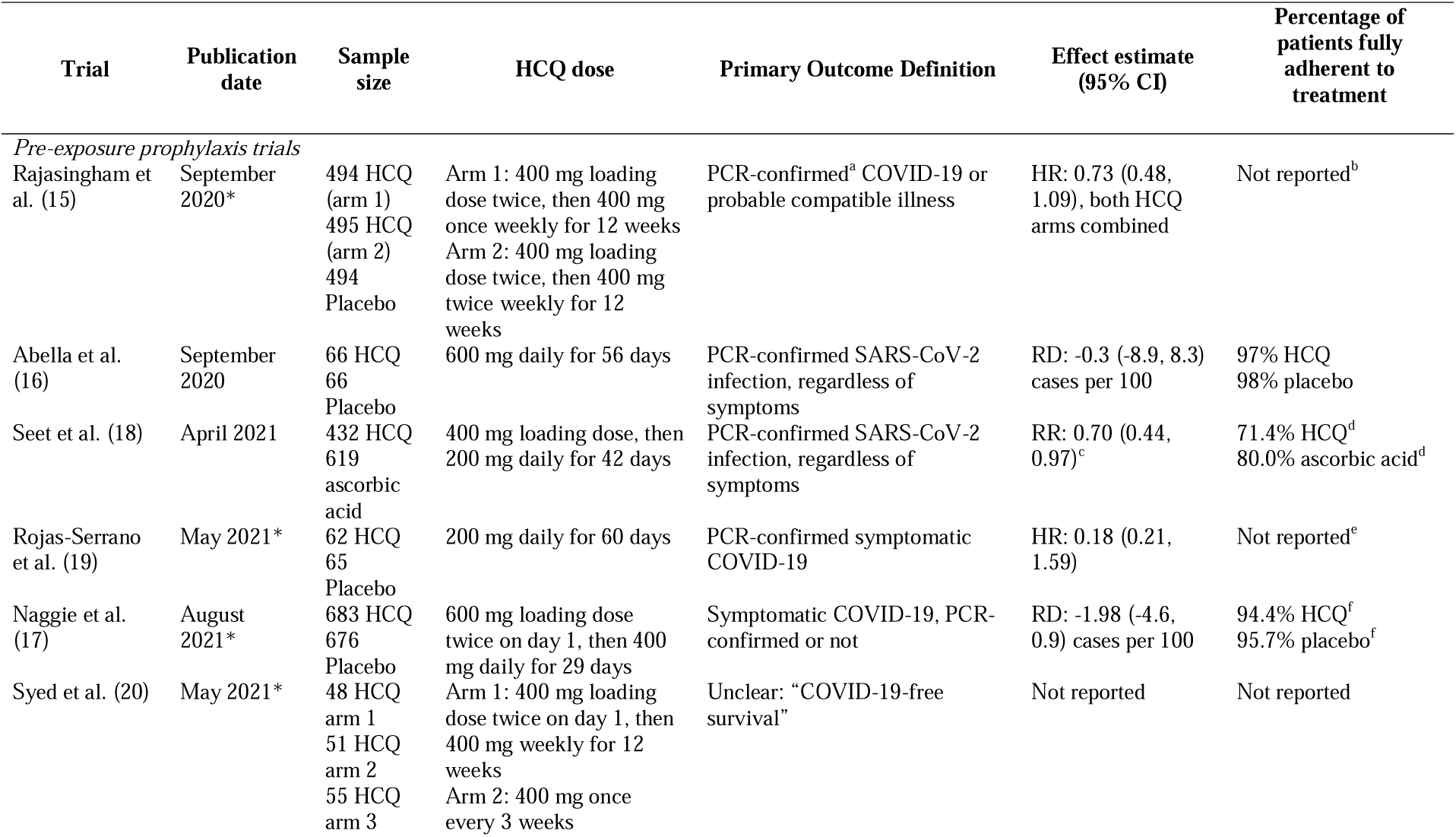
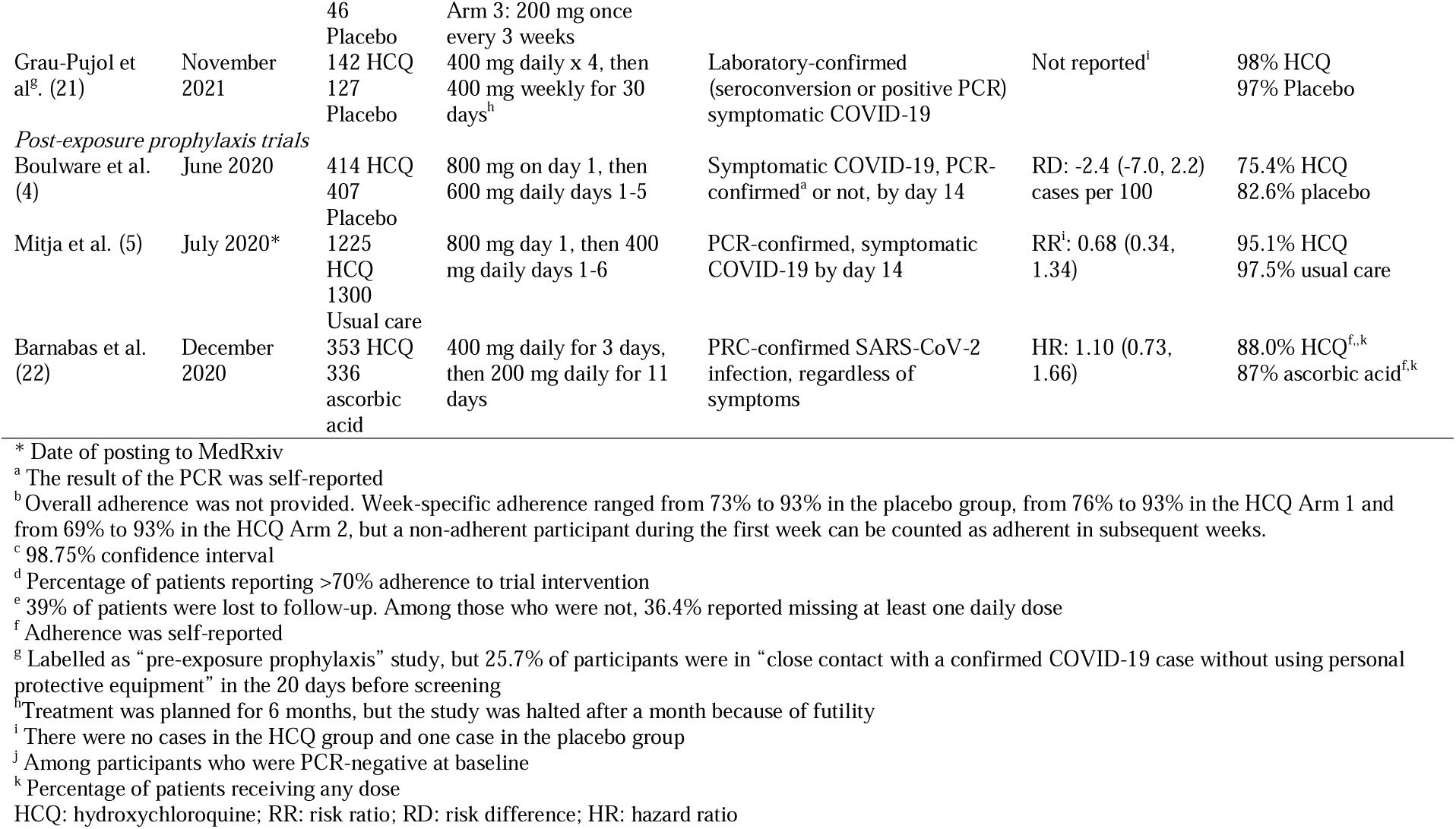
Summary of randomized trials studying hydroxychloroquine as prophylaxis for COVID-19

## Discussion

When considered together, the available pre-exposure prophylaxis randomized trials yield a point estimate of an approximately 30% lower risk of COVID-19 for assignment to HCQ compared with no HCQ among PCR-negative individuals at randomization. Any effect between approximately 48% and no reduction in risk is highly compatible with the data from these trials. The pooled effect estimate for the post-exposure randomized trial was closer to the null and both substantial reduction and moderate increase in risk were highly compatible with the data from the trials.

The choice of between common effect and random effects approaches for meta-analysis had little impact on the point estimates because the statistical heterogeneity across studies, as measured by the I^2^, was close to zero. There was some heterogeneity in the pre-exposure prophylaxis trials regarding the period over which the outcome was ascertained. However, the maximum difference was only 4 weeks and, given the long half-life of HCQ (25), variations in the order of weeks are not expected to substantially alter the trial estimates.

Our pooled estimates are based on the design and analytic choices made by the investigators of each trial. Ideally, these investigators could coordinate analyses of individual-level data with standardized outcome definitions; corrections for differences in length of follow-up and treatment dose; adjustment for losses to follow-up and other deviations from protocol (26); and adoption of a more causally-interpretable meta-analytic approach (27). All but one pre-exposure prophylaxis trials used laboratory-confirmed infection as the primary outcome; the remaining trial (15), and one of the three post-exposure prophylaxis trials (4), did not require laboratory confirmation and included some self-reported test results. Excluding this study resulted in a similar point estimate (Supplement Figure 2).

As the results of pre- and post-exposure prophylaxis trials appeared between June and September of 2020, the pooled point estimate hovered around a 20% reduction with increasingly narrower compatibility intervals (Supplementary Figure 4). Yet throughout this entire period the opinion of many medical researchers was that HCQ was ineffective for prevention and that additional trials were unnecessary. The point estimate moved to around 10% reduction after the publication in December of a post-exposure prophylaxis cluster-randomized study (22) in which the proportion of individuals with negative PCR at baseline differed between randomized groups and no outcomes were recorded among individuals with unavailable PCR at baseline. The practical implications of these baseline imbalances are unclear but may have guided the investigators’ decision to adjust for baseline variables, which was not contemplated in the study protocol (28).

The pooled estimates in Supplement Figure 4 combine pre- and post-exposure prophylaxis trials, under the assumption of a similar mechanism of action for both preventive approaches (25). However, the long time from exposure to treatment initiation in the post-exposure trials (several days as opposed to the 48 hours recommended for other viruses like HIV) weakens the case for pooling studies of pre- and post-exposure prophylaxis because the latter can resemble studies of early treatment. Therefore, as the number of completed randomized trials increased in 2021, meta-analyses like ours need to present separate estimates for pre-exposure and post-exposure prophylaxis.

This systematic review offers an important lesson for future research on drug repurposing: Recruitment for most trials of HCQ prophylaxis was severely impeded by incorrect interpretations of the evidence from the early (mostly post-exposure prophylaxis) trials. The findings from the first reported trials were widely (and incorrectly) portrayed as definite evidence of the lack of effectiveness of HCQ, simply because they were not “statistically significant” when taken individually. Thus, the common confusion between the concepts “no effect” and “no statistical significance” led many to prematurely conclude that HCQ had no prophylactic effect when the correct conclusion was that the effect estimate was still too imprecise (29, 30). We encourage the authors of living systematic reviews to update their findings frequently when, as in this case, public opinion interferes with the generation of the very evidence that will feed their systematic review.

In summary, throughout much of the pre-vaccine period of the pandemic, the available evidence was compatible with HCQ being viable as prophylaxis. Yet an incorrect interpretation of early inconclusive studies interfered with the timely completion of the trials needed to make a final determination. Though the availability of effective COVID-19 vaccines reduces the need for pharmacological prophylaxis, it is important to improve the process by which the medical community develops and interprets data before the next public health emergency arrives. To avoid he proliferation of small studies with different methodologies, national regulators and international health organizations can play a key role in the coordination and harmonization of the design of the randomized trials that they approve or endorse.

## Data Availability

Data used in the analysis are publicly available

## Acknowledgments

We thank Sander Greenland and Daniel Westreich for technical advice, Dr. Matthew Cefalu for constructive feedback on study outcomes, and Prof. Richard Riley, Dr. Tim Morrison, and Mr. David Fisher for constructive feedback on estimation methods for random effects meta-analysis. We thank Dr. Ruanne Barnabas and Torin Schaafsma for providing the risk ratio from their study for inclusion in this meta-analysis.

## References

1. Pan H, Peto R, Karim QA, Alejandria M, Henao-Restrepo AM, García CH, et al. Repurposed antiviral drugs for COVID-19 –interim WHO SOLIDARITY trial results. medRxiv. 2020:2020.10.15.20209817.

2. Horby P, Mafham M, Linsell L, Bell JL, Staplin N, Emberson JR, et al. Effect of Hydroxychloroquine in Hospitalized Patients with Covid-19. The New England journal of medicine. 2020;383(21):2030–40.

3. Bienvenu AL, Marty AM, Jones MK, Picot S. Systematic review of registered trials of Hydroxychloroquine prophylaxis for COVID-19 health-care workers at the first third of 2020. One health (Amsterdam, Netherlands). 2020;10:100141.

4. Boulware DR, Pullen MF, Bangdiwala AS, Pastick KA, Lofgren SM, Okafor EC, et al. A Randomized Trial of Hydroxychloroquine as Postexposure Prophylaxis for Covid-19. New England Journal of Medicine. 2020;383(6):517–25.

5. Mitjà O, Corbacho-Monné M, Ubals M, Alemany A, Suñer C, Tebé C, et al. A Cluster-Randomized Trial of Hydroxychloroquine for Prevention of Covid-19. New England Journal of Medicine. 2020.

6. Goldman JD. Hydroxychloroquine for Prevention of Severe Acute Respiratory Syndrome Coronavirus 2 (SARS-CoV-2) Infection: Challenges to Trial Conduct During the Global Pandemic. Clinical infectious diseases : an official publication of the Infectious Diseases Society of America. 2021;72(11):e844–e7.

7. Sterne JAC, Savović J, Page MJ, Elbers RG, Blencowe NS, Boutron I, et al. RoB 2: a revised tool for assessing risk of bias in randomised trials. BMJ. 2019;366:l4898.

8. DerSimonian R, Laird N. Meta-analysis in clinical trials. Controlled clinical trials. 1986;7(3):177–88.

9. Hartung J, Knapp G. A refined method for the meta-analysis of controlled clinical trials with binary outcome. Statistics in medicine. 2001;20(24):3875–89.

10. IntHout J, Ioannidis JPA, Borm GF. The Hartung-Knapp-Sidik-Jonkman method for random effects meta-analysis is straightforward and considerably outperforms the standard DerSimonian-Laird method. BMC Medical Research Methodology. 2014;14(1):25.

11. Wiksten A, Rücker G, Schwarzer G. Hartung-Knapp method is not always conservative compared with fixed-effect meta-analysis. Statistics in medicine. 2016;35(15):2503–15.

12. Jackson D, Law M, Rücker G, Schwarzer G. The Hartung-Knapp modification for random-effects meta-analysis: A useful refinement but are there any residual concerns? Statistics in medicine. 2017;36(25):3923–34.

13. Röver C, Knapp G, Friede T. Hartung-Knapp-Sidik-Jonkman approach and its modification for random-effects meta-analysis with few studies. BMC Medical Research Methodology. 2015;15(1):99.

14. Partlett C, Riley RD. Random effects meta-analysis: Coverage performance of 95% confidence and prediction intervals following REML estimation. Statistics in medicine. 2017;36(2):301–17.

15. Rajasingham R, Bangdiwala AS, Nicol MR, Skipper CP, Pastick KA, Axelrod ML, et al. Hydroxychloroquine as pre-exposure prophylaxis for COVID-19 in healthcare workers: a randomized trial. Clinical infectious diseases : an official publication of the Infectious Diseases Society of America. 2020.

16. Abella BS, Jolkovsky EL, Biney BT, Uspal JE, Hyman MC, Frank I, et al. Efficacy and Safety of Hydroxychloroquine vs Placebo for Pre-exposure SARS-CoV-2 Prophylaxis Among Health Care Workers: A Randomized Clinical Trial. JAMA internal medicine. 2020.

17. Naggie S, Milstone A, Castro M, Collins SP, Seetha L, Anderson DJ, et al. Hydroxychloroquine for pre-exposure prophylaxis of COVID-19 in health care workers: a randomized, multicenter, placebo-controlled trial (HERO-HCQ). medRxiv. 2021:2021.08.19.21262275.

18. Seet RCS, Quek AML, Ooi DSQ, Sengupta S, Lakshminarasappa SR, Koo CY, et al. Positive impact of oral hydroxychloroquine and povidone-iodine throat spray for COVID-19 prophylaxis: An open-label randomized trial. Int J Infect Dis. 2021;106:314–22.

19. Rojas-Serrano J, Thirion-Romero AMP-VI, Vázquez-Pérez J, Ramírez-Venegas FM-NA, Pérez-Kawabe KM, Pérez-Padilla R. Hydroxychloroquine For Prophylaxis Of COVID-19 In Health Workers: A Randomized Clinical Trial. Plos One. 2021:Upcoming.

20. Syed F, Arif MA, Niazi R, Baqar JB, Hashmi UL, Batool S, et al. Pre-Exposure Prophylaxis with Various Doses of Hdroxychloroquine among high-risk COVID 19 Healthcare Personnel: CHEER randomized controlled trial. medRxiv. 2021:2021.05.17.21257012.

21. Grau-Pujol B, Camprubí-Ferrer D, Marti-Soler H, Fernández-Pardos M, Carreras-Abad C, Andrés MV-d, et al. Pre-exposure prophylaxis with hydroxychloroquine for COVID-19: a double-blind, placebo-controlled randomized clinical trial. Trials. 2021;22(1):808.

22. Barnabas RV, Brown ER, Bershteyn A, Stankiewicz Karita HC, Johnston C, Thorpe LE, et al. Hydroxychloroquine as Postexposure Prophylaxis to Prevent Severe Acute Respiratory Syndrome Coronavirus 2 Infection : A Randomized Trial. Ann Intern Med. 2020.

23. Kuhar DT, Henderson DK, Struble KA, Heneine W, Thomas V, Cheever LW, et al. Updated U.S. Public Health Service guidelines for the management of occupational exposures to HIV and recommendations for postexposure prophylaxis. [Consensus Document on post-exposure prophylaxis against HIV, HBV and HCV in adults and children]. Enfermedades infecciosas y microbiologia clinica. 2016;34(2):121.e1-15.

24. Yao X, Ye F, Zhang M, Cui C, Huang B, Niu P, et al. In Vitro Antiviral Activity and Projection of Optimized Dosing Design of Hydroxychloroquine for the Treatment of Severe Acute Respiratory Syndrome Coronavirus 2 (SARS-CoV-2). Clinical infectious diseases : an official publication of the Infectious Diseases Society of America. 2020;71(15):732–9.

25. Hernan MA, Robins JM. Per-protocol analyses of pragmatic trials. N Engl J Med. 2017;377(14):1391–8.

26. Dahabreh IJ, Petito LC, Robertson SE, Hernán MA, Steingrimsson JA. Toward Causally Interpretable Meta-analysis: Transporting Inferences from Multiple Randomized Trials to a New Target Population. Epidemiology (Cambridge, Mass). 2020;31(3):334–44.

27. Barnabas RV, Brown E, Bershteyn A, Miller RS, Wener M, Celum C, et al. Efficacy of hydroxychloroquine for post-exposure prophylaxis to prevent severe acute respiratory syndrome coronavirus 2 (SARS-CoV-2) infection among adults exposed to coronavirus disease (COVID-19): a structured summary of a study protocol for a randomised controlled trial. Trials. 2020;21(1):475.

28. Amrhein V, Greenland S, McShane B. Scientists rise up against statistical significance. Nature. 2019;567(7748):305–7.

29. Greenland S, Senn SJ, Rothman KJ, Carlin JB, Poole C, Goodman SN, et al. Statistical tests, P values, confidence intervals, and power: a guide to misinterpretations. European journal of epidemiology. 2016;31(4):337–50.

