## Supplementary Material for "Systematic review and meta-analysis of randomized trials of hydroxychloroquine for the prevention of COVID-19"

January 25, 2022

**hydroxychloroquine for the prevention of COVID-19**

Xabier García-Albéniz^1,2^, Julia del Amo^3^, Rosa Polo^3^,

José Miguel Morales-Asencio^4^, Miguel A Hernán^2,5^

^1^ RTI Health Solutions. Barcelona, Spain

^2^ CAUSALab. Harvard T.H. Chan School of Public Health. Boston, MA, USA

^3^ National Plan Against AIDS, Ministry of Health. Madrid, Spain

^4^ Department of Nursing and Podiatry, Universidad de Málaga, Málaga, Spain.

^5^ Departments of Epidemiology and Biostatistics, Harvard T.H. Chan School of Public Health, Boston, MA, USA; Harvard-MIT Division of Health Sciences and Technology, Boston, MA, USA

**Supplementary Material**

**Supplementary Figure 1.** Risk difference estimates (cases per 100) of COVID-19 for hydroxychloroquine vs. no hydroxychloroquine in randomized trials of pre-exposure prophylaxis, pooled and by study. These estimates are based on the definition of COVID-19 reported in the primary analysis of each study.

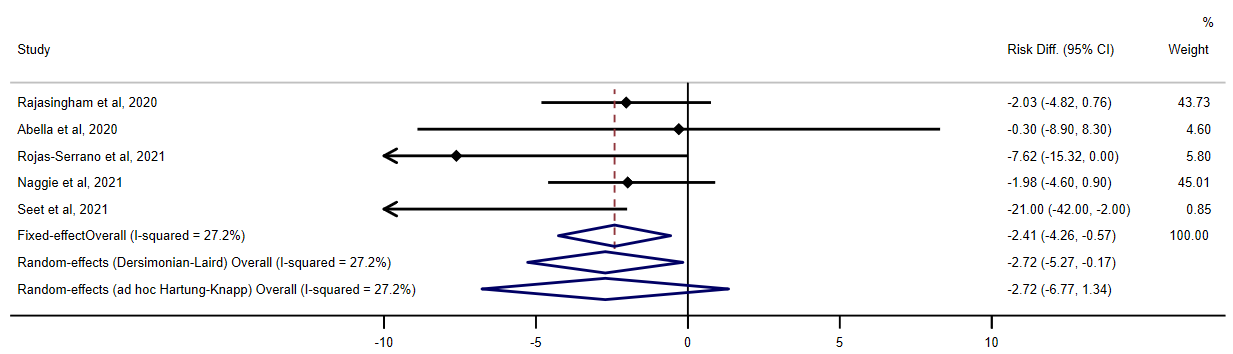

**Supplementary Figure 2**. Risk ratio estimates of “laboratory-confirmed, symptomatic COVID-19” for hydroxychloroquine vs. no hydroxychloroquine in randomized trials of pre-exposure prophylaxis, pooled and by study.

Note: The estimates for the studies by Rajasingham et al. and by Naggie et al. are based on a nonrandom subset of the cases that should have been laboratory-confirmed because participants in these studies had limited access to testing.

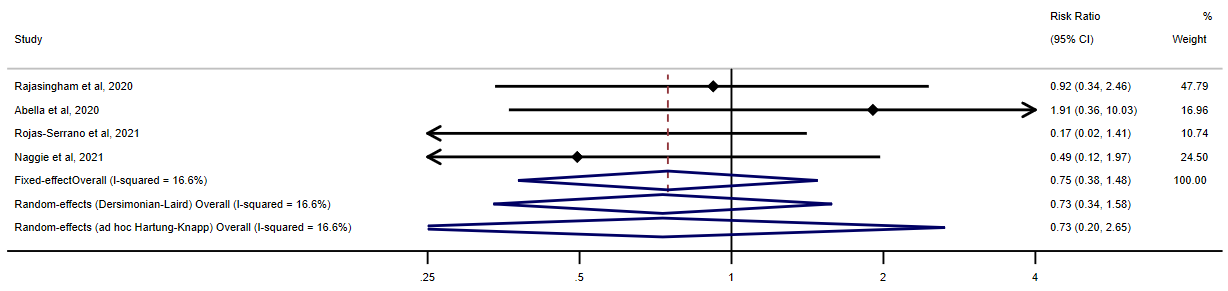

**Supplementary Figure 3.** Risk ratio estimates of COVID-19 for hydroxychloroquine vs. no hydroxychloroquine in randomized trials of pre-exposure prophylaxis published in peer-reviewed journals, pooled and by study. These estimates are based on the definition of COVID-19 reported in the primary analysis of each study.

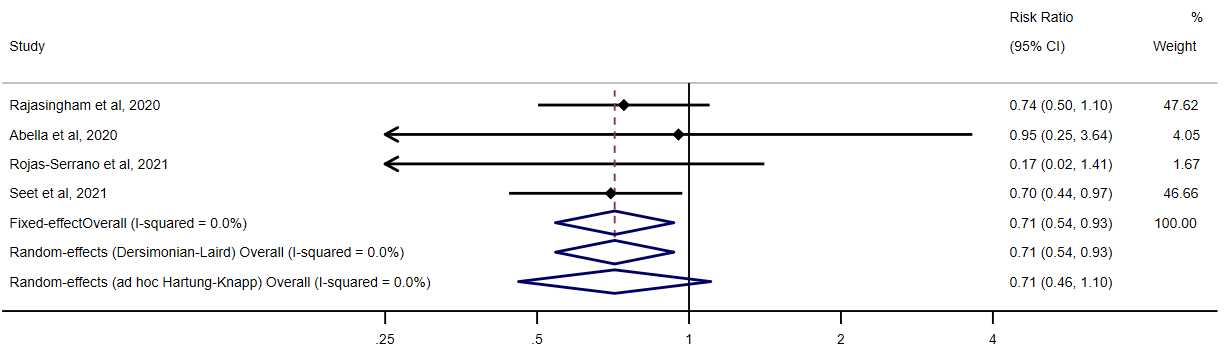

**Supplementary Figure 4.** Risk ratio estimates of COVID-19 (as reported in the primary analysis of each study) for hydroxychloroquine vs. no hydroxychloroquine in randomized trials of pre- and post-exposure prophylaxis, pooled as evidence gathered over time. Confidence intervals are computed using the standard random effects method.

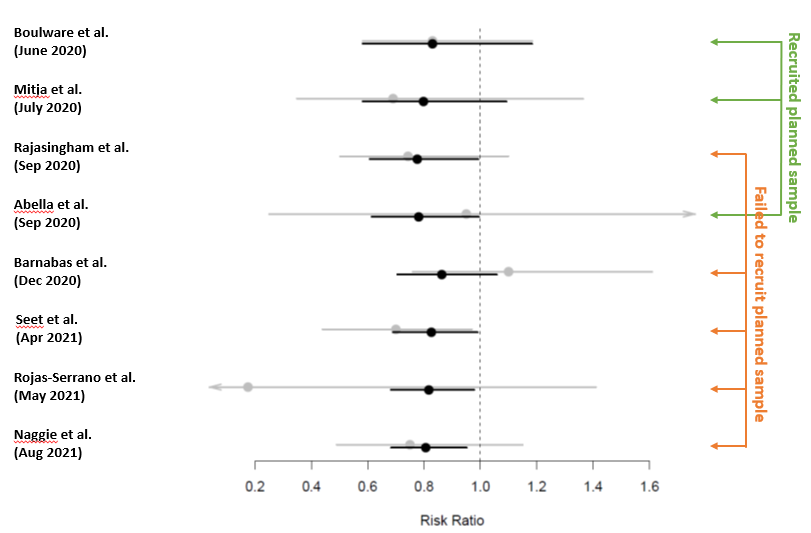

**PubMed search strategy:**

("hydroxychloroquine"[MeSH Terms] OR "hydroxychloroquine"[All Fields]) AND (("Pre-Exposure Prophylaxis"[Mesh]) OR "Post-Exposure Prophylaxis"[Mesh] OR prophylaxis[Title]) AND (("severe acute respiratory syndrome coronavirus 2"[Supplementary Concept] OR "severe acute respiratory syndrome coronavirus 2"[All Fields] OR "SARS-CoV-2"[All Fields]) AND ("COVID-19"[All Fields] OR "COVID-2019"[All Fields] OR "severe acute respiratory syndrome coronavirus 2"[Supplementary Concept] OR "severe acute respiratory syndrome coronavirus 2"[All Fields] OR "2019-nCoV"[All Fields] OR "SARS-CoV-2"[All Fields] OR "2019nCoV"[All Fields] OR ("Wuhan"[All Fields] AND ("coronavirus"[MeSH Terms] OR "coronavirus"[All Fields]) AND (2019/12/01:2019/12/31[Date - Publication] OR 2020/01/01:2020/12/31[Date - Publication])))) AND ("Clinical Trial"[Publication Type] OR "Clinical Trials as Topic"[MeSH Terms] OR "trial*"[Title]) NOT (“Review”[Publication Type] OR “Systematic review”[Publication Type]) NOT (“Animals”[Mesh] NOT “Humans”[Mesh])

**Risf of Bias**

Cochrane risk of bias 2.0 tool assessment of randomized trials

|  | Rajasingham et al., 2020 | Abella et al., 2020 | Boulware et al., 2020 | Mitja et al., 2020 | Barnabas et al., 2020 | Naggie et al., 2021 | Rojas-Serrano et al., 2021 | Seet et al., 2021 | Syed et al., 2021 | Grau-pujol et al., 2021 |
| --- | --- | --- | --- | --- | --- | --- | --- | --- | --- | --- |
| 1.1 Was the allocation sequence random? | Y | Y | Y | Y | Y | Y | Y | Y | Y | Y |
| 1.2 Was the allocation sequence concealed until participants were recruited and assigned to interventions? | Y | Y | Y | Y | Y | Y | Y | Y | NI | Y |
| 1.3 Did baseline differences between intervention groups suggest a problem with the randomization process? | N | N | N | N | N^1^ | N | N | N | PN | N |
| Risk of bias judgement | Low | Low | Low | Low | Low | Low | Low | Low | Some concerns | Low |
| 2.1. Were participants aware of their assigned intervention during the trial? | N | N | N | Y | N | N | N | Y | NI | N |
| 2.2. Were carers and people delivering the interventions aware of participants' assigned intervention during the trial? | N | N | N | Y | N | N | N | Y | NI | N |
| 2.3. If Y/PY/NI to 2.1 or 2.2: Were there deviations from the intended intervention that arose because of the trial context? |  |  |  | Y |  |  |  | N | N |  |
| 2.4 If Y/PY to 2.3: Were these deviations likely to have affected the outcome? |  |  |  | Y |  |  |  |  |  |  |
| 2.5. If Y/PY/NI to 2.4: Were these deviations from intended intervention balanced between groups? |  |  |  | N |  |  |  |  |  |  |
| 2.6 Was an appropriate analysis used to estimate the effect of assignment to intervention? | N^2^ | N^3^ | N^4^ | N^5^ | N^6^ | Y | Y | Y | N^7^ | N^9^ |
| 2.7 If N/PN/NI to 2.6: Was there potential for a substantial impact (on the result) of the failure to analyze participants in the group to which they were randomized? | Y | Y | Y | Y | Y |  |  | N | N | Y^9^ |
| Risk-of-bias judgement | Some concerns | Some concerns | Some concerns | Some concerns | Some concerns | Low | Low | Low | Some concerns | High |
| 3.1 Were data for this outcome available for all, or nearly all, participants randomized? | PN | Y | N | N | Y | Y | Y | Y | NI^8^ | Y |
| 3.2 If N/PN/NI to 3.1: Is there evidence that the result was not biased by missing outcome data? | N |  | N | N |  |  |  |  | N |  |
| 3.3 If N/PN to 3.2: Could missingness in the outcome depend on its true value? | Y |  | NI | Y |  |  |  |  | Y |  |
| 3.4 If Y/PY/NI to 3.3: Is it likely that missingness in the outcome depended on its true value? | NI |  | NI | NI |  |  |  |  | NI |  |
| Risk-of-bias judgement | Some concerns | Low | Some concerns | Some concerns | Low | Low | Low | Low | Some concerns | Low |
| 4.1 Was the method of measuring the outcome inappropriate? | Y | Y | Y | Y | N | Y | Y | N | Y | Y |
| 4.2 Could measurement or ascertainment of the outcome have differed between intervention groups? | N | N | N | N | N | N |  | N | PN | N |
| 4.3 If N/PN/NI to 4.1 and 4.2: Were outcome assessors aware of the intervention received by study participants? |  |  |  |  |  |  |  |  | NI |  |
| 4.4 If Y/PY/NI to 4.3: Could assessment of the outcome have been influenced by knowledge of intervention received? |  |  |  |  |  |  |  |  | NI |  |
| 4.5 If Y/PY/NI to 4.4: Is it likely that assessment of the outcome was influenced by knowledge of intervention received? |  |  |  |  |  |  |  |  | PN |  |
| Risk-of-bias judgement | Low | Low | Low | Low | Low | Low | Low | Low | Some concerns | Low |
| 5.1 Were the data that produced this result analysed in accordance with a pre-specified analysis plan that was finalized before unblinded outcome data were available for analysis? | Y | Y | Y | NI | N^5^ | Y | Y | NI | NI | Y |
| Is the numerical result being assessed likely to have been selected, on the basis of the results, from... |  |  |  |  |  |  |  |  |  |  |
| 5.2. ... multiple eligible outcome measurements (e.g. scales, definitions, time points) within the outcome domain? | N | N | N | N | N | N | N | N | N | N |
| 5.3 ... multiple eligible analyses of the data? | N | N | N | N | N | N | N | N | N | N |
| Risk-of-bias judgement | Low | Low | Low | Low | Low | Low | Low | Low | Low | Low |

Y: Yes; PY: Probably Yes; PN: Probably No; N: No; NI: No Information

^1^ The proportion of patients excluded for not having a negative PCR at baseline differed by exposure arm: 13.3% in the HCQ arm and 20.4% in the control group (95% CI of the risk difference is -12.2% to -2.0%). No cases were detected among those with an inconclusive PCR at baseline (13.4% were expected if the risk was the same as for those with a negative PCR at baseline).

^2^ Thirteen patients were excluded after randomization for reaching the primary endpoint before starting the study drug.

^3^ Seven patients were excluded after randomization for presenting a positive test at baseline, never took the study medication or withdrew from the study.

^4^ One hundred patients were excluded after randomization for presenting symptoms at the time of receiving the trial intervention. Over 10% of the remaining patients did not have complete information on the outcome because they did not complete the follow-up.

^5^ Over 7% of randomized patients were excluded from the analysis for not having complete outcome data.

^6^ Adjustment for age, sex, and quarantine status was not specified in the study protocol (1).

^7^ No effect was estimated. Maximum disease severity (the likely primary outcome) was compared with an ANOVA test

^8^ A definition of the outcome is not provided

^9^ No effect was estimated on the basis of absence of events in the HCQ group. The outcomes were counted among participants who did not discontinue the intervention after randomization. Reasons for discontinuing the intervention included SARS-CoV-2 infection.

1.	Barnabas RV, Brown E, Bershteyn A, Miller RS, Wener M, Celum C, et al. Efficacy of hydroxychloroquine for post-exposure prophylaxis to prevent severe acute respiratory syndrome coronavirus 2 (SARS-CoV-2) infection among adults exposed to coronavirus disease (COVID-19): a structured summary of a study protocol for a randomised controlled trial. Trials. 2020;21(1):475.
